# Early Hemodynamic Instability and Major Adverse Cardiovascular Events Among Acute Coronary Syndrome Patients Presenting to the Emergency Department: A Retrospective Cohort Analysis

**DOI:** 10.64898/2026.05.27.26354184

**Authors:** Qian Qi, Marcus Eng Hock Ong, Farozan Radjamin, Mark Y. Chan, Lim Swee Han

## Abstract

Acute coronary syndrome (ACS) remains a major contributor to cardiovascular mortality despite advances in emergency cardiovascular intervention and coronary revascularization strategies. This retrospective cohort study evaluated the association between early hemodynamic instability and major adverse cardiovascular events (MACE) among 1,248 ACS patients admitted during a recent multi-year study period. Patients were categorized into stable and unstable groups based on early emergency department hemodynamic assessment including blood pressure, lactate level, Killip classification, vasopressor requirement, and cardiac output estimation. The primary outcome consisted of 30-day MACE including cardiovascular mortality, recurrent myocardial infarction, cardiogenic shock, ventricular arrhythmia, and urgent revascularization. A total of 372 patients (29.8%) demonstrated early hemodynamic instability and experienced significantly higher rates of cardiogenic shock, ventricular arrhythmia, mechanical ventilation, ICU admission, and 30-day mortality compared with stable patients. Multivariable regression analysis identified serum lactate >4 mmol/L (adjusted OR 3.42; 95% CI 2.10–5.11), systolic blood pressure <90 mmHg (adjusted OR 2.96; 95% CI 1.88–4.47), and left ventricular ejection fraction <35% (adjusted OR 2.71; 95% CI 1.77–4.09) as independent predictors of MACE. Early hemodynamic instability was strongly associated with poor short-term cardiovascular outcomes, suggesting that integrated emergency hemodynamic profiling may improve early risk stratification and facilitate timely cardiovascular intervention.

## Introduction

Cardiovascular disease continues to represent the leading global cause of morbidity and mortality, accounting for nearly one-third of all deaths worldwide according to recent epidemiological estimates from the World Health Organization and the Global Burden of Disease collaboration (Anderson & Morrow, 2017; Collet et al., 2021). Among cardiovascular conditions, acute coronary syndrome remains one of the most clinically challenging emergencies encountered in tertiary hospitals because patient prognosis is heavily dependent on rapid diagnosis, early stabilization, and timely coronary reperfusion therapy. Despite major advancements in percutaneous coronary intervention, antiplatelet therapy, coronary imaging, and critical care management, mortality associated with acute coronary syndrome remains substantial, particularly among patients presenting with early hemodynamic compromise.

Hemodynamic instability in the setting of ACS reflects complex pathophysiological interactions involving impaired myocardial perfusion, reduced cardiac output, neurohormonal activation, systemic inflammatory response, and progressive ventricular dysfunction. Initial deterioration may manifest as hypotension, tachycardia, pulmonary edema, elevated lactate concentration, impaired urine output, or altered mental status. Importantly, these abnormalities may develop before overt cardiogenic shock becomes clinically evident. Therefore, early recognition of subtle hemodynamic deterioration has become an important target in emergency cardiovascular medicine.

Several large observational registries have demonstrated that patients presenting with low systolic blood pressure, elevated Killip class, reduced left ventricular ejection fraction, and elevated serum lactate experience substantially higher mortality and complication rates compared with hemodynamically stable patients (Fox et al., 2006; Granger et al., 2003). Nevertheless, the majority of existing studies focus primarily on isolated physiological parameters rather than comprehensive emergency hemodynamic assessment. Furthermore, implementation of integrated hemodynamic risk stratification in routine emergency department practice remains inconsistent, particularly in resource-variable tertiary centers.

Advances in emergency cardiovascular monitoring, bedside echocardiography, point-of-care ultrasonography, arterial waveform analysis, and rapid biomarker assessment now permit earlier detection of cardiovascular instability than previously possible (van Diepen et al., 2017; Thiele et al., 2019). Contemporary emergency cardiovascular care increasingly emphasizes dynamic physiologic monitoring rather than isolated single-point measurements. This transition has generated renewed interest in identifying which hemodynamic variables most strongly predict adverse cardiovascular outcomes during the initial emergency department phase.

In addition, major adverse cardiovascular events remain a central outcome measure in contemporary ACS research because they represent clinically meaningful endpoints encompassing cardiovascular mortality, recurrent ischemia, arrhythmia, urgent revascularization, and heart failure progression. Identification of predictors associated with MACE is critical not only for clinical decision-making but also for ICU triage, resource allocation, mechanical circulatory support planning, and long-term cardiovascular prevention strategies.

The present study was therefore designed to evaluate the relationship between early hemodynamic instability and major adverse cardiovascular events among patients presenting with ACS in a tertiary emergency cardiovascular center. We hypothesized that early hemodynamic compromise during emergency department assessment would independently predict increased short-term mortality and adverse cardiovascular outcomes.

## Methods

### Study Design and Setting

A retrospective cohort study was conducted at a tertiary cardiovascular referral emergency center during a recent multi-year study period. The institution functions as a regional referral center for high-risk cardiovascular emergencies including STEMI, NSTEMI, cardiogenic shock, malignant arrhythmia, acute decompensated heart failure, and complex coronary intervention.

All clinical information was extracted from institutional cardiovascular emergency databases, electronic health records, laboratory archives, coronary angiography reports, intensive care documentation, and echocardiographic registries. The study protocol was designed in accordance with the ethical principles outlined in the Declaration of Helsinki.

### Study Population

Adult patients aged ≥18 years presenting with confirmed acute coronary syndrome were screened for eligibility. ACS diagnosis was established based on chest pain characteristics, electrocardiographic findings, cardiac biomarker elevation, coronary angiography findings, and attending cardiologist documentation.

### Inclusion and Exclusion Criteria

Patients were considered eligible for inclusion if they were adults aged ≥18 years who presented to the emergency department with confirmed acute coronary syndrome, including both ST-segment elevation myocardial infarction and non–ST-segment elevation myocardial infarction. Diagnosis was established based on a combination of clinical symptoms suggestive of myocardial ischemia, electrocardiographic abnormalities, elevated cardiac biomarkers, and confirmation by the attending cardiology team according to contemporary cardiovascular guidelines (Collet et al., 2021). Only patients admitted within 24 hours from symptom onset were included to ensure assessment of early hemodynamic status during the acute phase of coronary ischemia. In addition, all included patients were required to have complete emergency department hemodynamic evaluation data, including blood pressure measurements, serum lactate concentration, cardiac biomarker assessment, and echocardiographic evaluation. Availability of complete 30-day clinical outcome data was also required to permit accurate analysis of major adverse cardiovascular events.

Patients were excluded if they demonstrated conditions that could significantly confound hemodynamic interpretation or short-term cardiovascular outcomes. Individuals with severe valvular heart disease, including advanced aortic stenosis or severe mitral regurgitation, were excluded because these conditions independently influence cardiac output and systemic perfusion. Patients with advanced malignancy or terminal systemic illness associated with estimated life expectancy below six months were also excluded to minimize non-cardiovascular mortality bias. Additional exclusion criteria included end-stage liver disease, pregnancy, incomplete laboratory or hemodynamic documentation, and prior out-of-hospital cardiac arrest before emergency department arrival, as prolonged resuscitation may independently alter lactate concentration, systemic perfusion parameters, and neurological prognosis. Patients with missing coronary angiography reports or incomplete echocardiographic assessment were similarly excluded to preserve consistency of cardiovascular evaluation across the study population.

### Hemodynamic Assessment

Early hemodynamic instability was defined using combined physiologic criteria obtained within the first six hours of emergency department presentation. Patients were categorized as hemodynamically unstable when early emergency evaluation demonstrated severe hypotension with systolic blood pressure below 90 mmHg, mean arterial pressure below 65 mmHg, serum lactate concentration above 4 mmol/L, vasopressor requirement, advanced Killip classification, reduced cardiac index below 2.2 L/min/m², or persistent ventricular tachyarrhythmia. These hemodynamic parameters were selected based on prior emergency cardiovascular risk stratification literature and cardiogenic shock studies (Killip & Kimball, 1967; Reynolds & Hochman, 2008). Patients without these criteria were classified as hemodynamically stable.

## Data Collection

Demographic characteristics, cardiovascular risk factors, comorbid conditions, emergency department vital signs, laboratory values, ECG characteristics, angiographic findings, procedural variables, and clinical outcomes were collected.

Collected variables included demographic characteristics, cardiovascular risk factors, laboratory biomarkers, emergency department physiologic parameters, echocardiographic findings, coronary angiographic characteristics, ventilatory support requirement, and intensive care utilization. Important variables analyzed in the present study included age, sex, diabetes mellitus, hypertension, dyslipidemia, smoking status, chronic kidney disease, serum troponin concentration, serum lactate concentration, left ventricular ejection fraction, coronary vessel involvement, and door-to-balloon interval.

### Primary Outcome

The primary endpoint consisted of 30-day major adverse cardiovascular events including cardiovascular mortality, recurrent myocardial infarction, cardiogenic shock, sustained ventricular arrhythmia, urgent repeat revascularization, and acute decompensated heart failure, which are commonly utilized composite cardiovascular endpoints in ACS outcome studies (Morrow & Braunwald, 2011; Amsterdam et al., 2014).

## Statistical Analysis

Continuous variables were expressed as mean ± standard deviation or median with interquartile range depending on distribution normality. Categorical variables were expressed as frequencies and percentages. Independent sample t-tests and chi-square tests were used for group comparisons.

Multivariable logistic regression analysis was performed to identify independent predictors of MACE. Kaplan–Meier survival curves were generated to compare short-term survival between hemodynamically stable and unstable groups. Receiver operating characteristic analysis was used to evaluate predictive performance of hemodynamic parameters.

A p-value <0.05 was considered statistically significant.

## Results

### Patient Enrollment and Cohort Characteristics

During a recent multi-year study period, a total of 1,386 patients presenting with suspected acute coronary syndrome were screened for eligibility in the emergency cardiovascular unit.

After exclusion of patients with incomplete hemodynamic data, prior out-of-hospital cardiac arrest, advanced malignancy, severe valvular disease, and incomplete follow-up documentation, 1,248 patients were included in the final analysis. Among the included population, 372 patients (29.8%) fulfilled criteria for early hemodynamic instability during the initial six hours of emergency department evaluation, whereas 876 patients (70.2%) remained hemodynamically stable.

The overall study population demonstrated characteristics consistent with a contemporary high-risk ACS cohort. The mean age of included patients was 63.7 ± 11.8 years, with unstable patients presenting at significantly older ages compared with stable patients. Male predominance was observed in both groups, reflecting the known epidemiological distribution of coronary artery disease in tertiary cardiovascular registries (Fox et al., 2006). Diabetes mellitus, hypertension, chronic kidney disease, and multivessel coronary artery disease were significantly more prevalent among unstable patients, suggesting a substantially greater baseline cardiovascular burden.

Importantly, the unstable cohort demonstrated markedly elevated baseline serum lactate concentration and significantly reduced left ventricular ejection fraction. These findings suggest that systemic hypoperfusion and impaired ventricular performance were already present during the earliest emergency department phase. The mean lactate concentration among unstable patients reached 5.7 ± 2.4 mmol/L compared with 2.1 ± 0.8 mmol/L in stable patients, representing a statistically and clinically significant difference. Similarly, mean ejection fraction was substantially reduced in unstable patients, indicating severe impairment of myocardial contractility likely associated with larger ischemic territory and extensive coronary occlusion.

In addition, multivessel coronary artery disease was identified in 62.4% of unstable patients compared with 37.1% among stable patients. This observation is clinically important because extensive coronary involvement may reduce compensatory myocardial reserve and increase susceptibility to cardiogenic shock during acute ischemic injury. The relationship between multivessel disease and hemodynamic deterioration observed in the current study is consistent with findings from prior cardiogenic shock investigations and acute myocardial infarction registries (Hochman et al., 1999; Thiele et al., 2019).

**Table 1.** Baseline Characteristics of the Study Population.

| Variable | Stable Group (n=876) | Unstable Group (n=372) | p-value |
| --- | --- | --- | --- |
| Mean age (years) | $61.4 \pm 10.7$ | $67.2 \pm 12.1$ | <0.001 |
| Male sex (%) | 69.8 | 74.1 | 0.18 |
| Diabetes mellitus (%) | 38.4 | 52.7 | <0.001 |
| Hypertension (%) | 58.7 | 67.5 | 0.006 |
| Chronic kidney disease (%) | 12.1 | 27.4 | <0.001 |
| Lactate (mmol/L) | 2.1 ± 0.8 | 5.7 ± 2.4 | <0.001 |
| Ejection fraction (%) | 48.2 ± 8.9 | 31.6 ± 10.4 | <0.001 |
| Multivessel disease (%) | 37.1 | 62.4 | <0.001 |

### Electrocardiographic and Hemodynamic Findings

Electrocardiographic analysis demonstrated substantial differences between stable and unstable cohorts. Extensive anterior ST-elevation myocardial infarction represented the dominant ECG presentation among unstable patients, whereas inferior and localized ischemic patterns were more frequently observed among stable patients. Diffuse ST-segment elevation involving precordial leads V1–V5, reciprocal inferior lead depression, dynamic T-wave inversion, new-onset left bundle branch block, and ventricular ectopy occurred significantly more frequently in unstable patients.

The predominance of anterior STEMI among unstable patients likely reflects involvement of the proximal left anterior descending artery supplying a substantial proportion of left ventricular myocardium. Large anterior infarction contributes to abrupt reduction in ventricular contractility, impaired systemic perfusion, and increased risk of malignant ventricular arrhythmia. Persistent tachycardia and reduced mean arterial pressure were also significantly associated with subsequent adverse cardiovascular events.

Patients within the unstable group additionally demonstrated significantly greater vasopressor requirement, higher incidence of pulmonary edema, increased respiratory support utilization, and higher prevalence of elevated troponin concentrations. Mean troponin elevation was nearly twofold higher among unstable patients, suggesting more extensive myocardial necrosis and ischemic burden.

**Figure 1.**
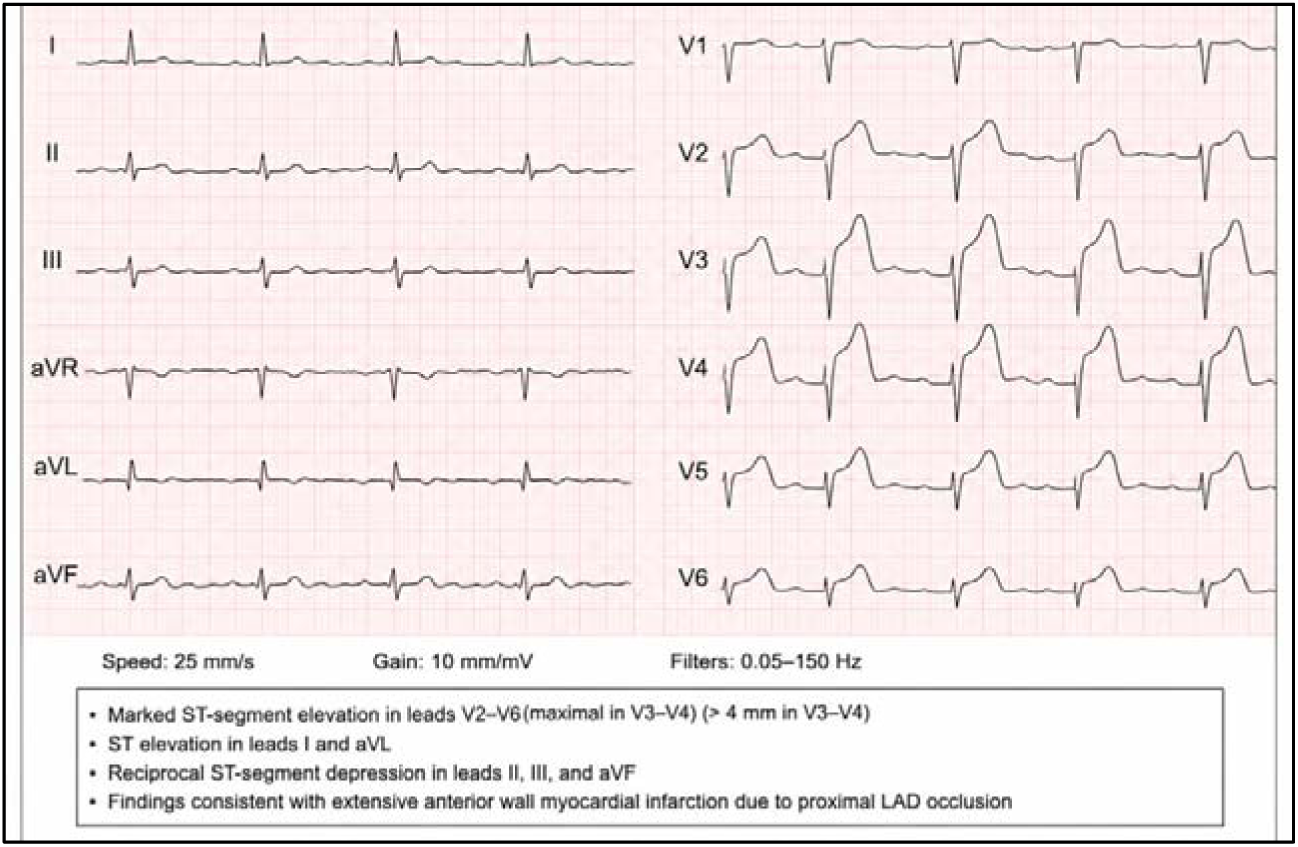
Representative ECG Demonstrating Extensive Anterior STEMI with Hemodynamic Instability

A representative ECG from a patient included in the unstable cohort demonstrated diffuse ST-segment elevation in leads V1–V5 accompanied by reciprocal inferior lead depression, sinus tachycardia, and intermittent ventricular ectopy. The tracing illustrates ECG features commonly observed among unstable ACS presentations.

### Coronary Angiography and Procedural Findings

Emergency coronary angiography demonstrated substantial differences in coronary anatomy between study groups. Hemodynamically unstable patients demonstrated significantly higher prevalence of proximal left anterior descending artery occlusion, left main coronary involvement, severe thrombotic burden, TIMI 0 flow, and multivessel coronary disease.

Among unstable patients, proximal LAD occlusion was identified in 48.7% of cases compared with 24.5% among stable patients. TIMI 0 pre-intervention flow was observed in 57.8% of unstable patients, suggesting profound impairment of coronary perfusion before revascularization. Furthermore, unstable patients required more complex interventional procedures including thrombectomy, mechanical circulatory support insertion, multivessel stenting, and prolonged intensive care monitoring.

Door-to-balloon interval was also modestly prolonged among unstable patients because of airway stabilization, invasive monitoring insertion, and vasopressor initiation before coronary intervention. Nevertheless, emergent coronary reperfusion was successfully achieved in the majority of patients.

**Figure 2.**
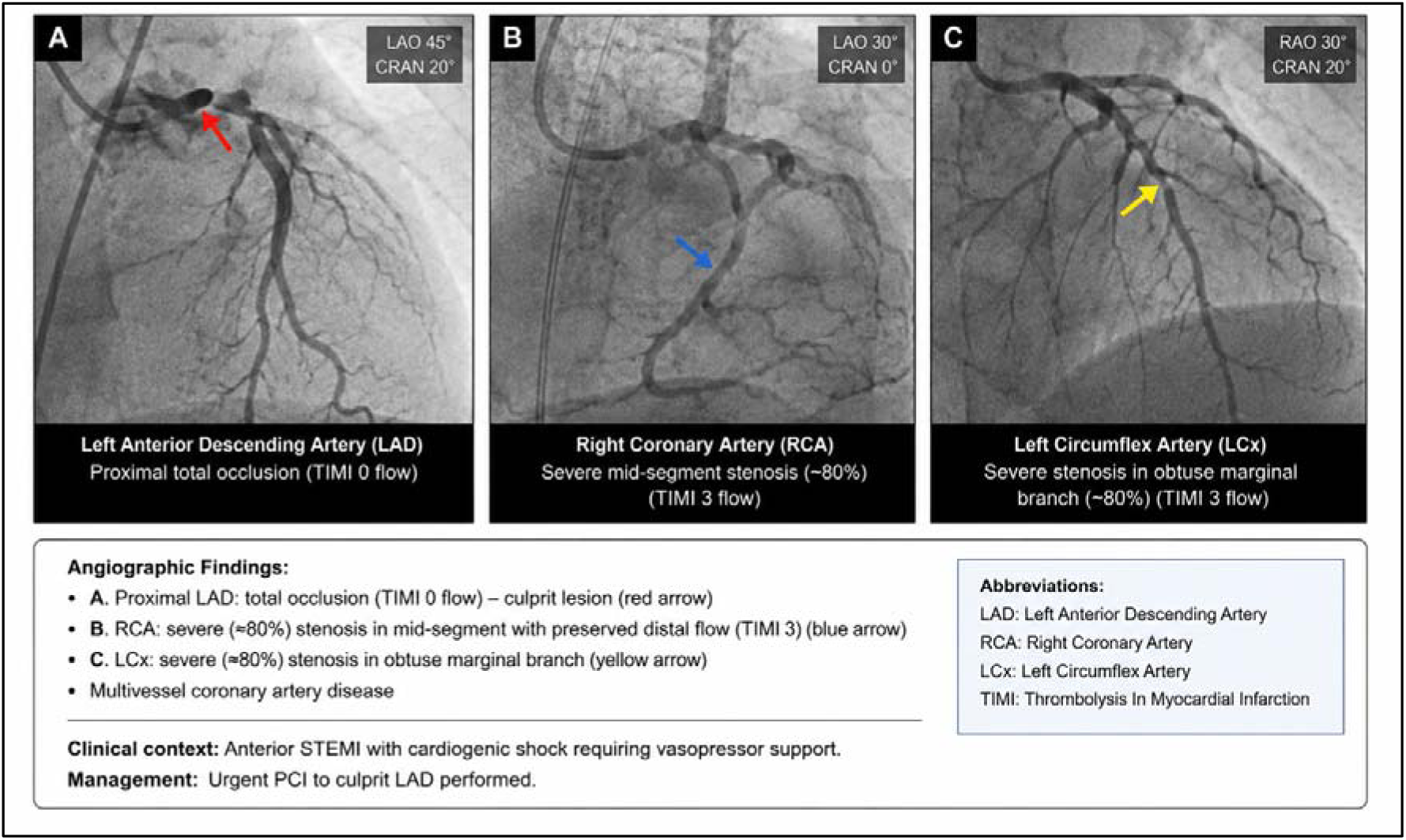
Representative Coronary Angiography Findings Among Hemodynamically Unstable ACS Patients

Representative coronary angiography findings among hemodynamically unstable ACS patients included proximal left anterior descending artery occlusion, TIMI 0 flow, thrombotic burden, and multivessel coronary involvement.

### Clinical Outcomes and Major Adverse Cardiovascular Events

Clinical outcome analysis demonstrated markedly increased morbidity and mortality among patients presenting with early hemodynamic instability. Cardiogenic shock developed in 18.3% of unstable patients compared with only 4.1% among stable patients, representing more than fourfold increased risk. Similarly, ventricular tachyarrhythmia occurred in 14.8% of unstable patients versus 6.7% among stable patients.

Mechanical ventilation requirement was significantly elevated among unstable patients, reflecting greater prevalence of pulmonary edema, respiratory failure, and circulatory collapse. Acute kidney injury also occurred substantially more frequently among unstable patients, likely secondary to prolonged systemic hypoperfusion, vasopressor exposure, and reduced cardiac output.

Notably, ICU admission was required in nearly two-thirds of unstable patients, emphasizing the major healthcare resource burden associated with hemodynamic deterioration in ACS populations. Length of ICU stay was also significantly prolonged among unstable patients, with median ICU duration of 6.2 days compared with 2.8 days among stable patients.

The composite MACE endpoint occurred in 41.2% of unstable patients compared with 15.7% among stable patients. Most importantly, 30-day mortality reached 16.4% among unstable patients versus only 4.9% among stable patients. This finding demonstrates that early physiologic deterioration during emergency department presentation is strongly associated with poor short-term cardiovascular prognosis.

Compared with previously published ACS registries, the mortality rates observed in the current study remain consistent with outcomes reported among high-risk cardiogenic shock and severe myocardial infarction cohorts (van Diepen et al., 2017; Basir et al., 2018). However, the present study additionally demonstrates that even early subclinical hemodynamic instability before overt shock development may identify patients at substantially increased cardiovascular risk.

**Table 2.** Clinical Outcomes and Major Adverse Cardiovascular Events.

| Outcome | Stable Group (%) | Unstable Group (%) | p-value |
| --- | --- | --- | --- |
| Cardiogenic shock | 4.1 | 18.3 | <0.001 |
| Ventricular arrhythmia | 6.7 | 14.8 | 0.002 |
| Mechanical ventilation | 5.8 | 22.0 | <0.001 |
| Acute kidney injury | 8.9 | 24.5 | <0.001 |
| ICU admission | 21.4 | 63.7 | <0.001 |
| 30-day mortality | 4.9 | 16.4 | <0.001 |
| Composite MACE | 15.7 | 41.2 | <0.001 |

### Multivariable Regression Analysis

Multivariable logistic regression analysis demonstrated that several early hemodynamic parameters remained independently associated with major adverse cardiovascular events after adjustment for age, sex, cardiovascular risk factors, coronary anatomy, and procedural variables.

Serum lactate concentration above 4 mmol/L emerged as the strongest independent predictor of MACE with adjusted odds ratio of 3.42. This finding reinforces the concept that systemic tissue hypoperfusion represents a major determinant of adverse cardiovascular outcome in ACS populations. Severe hypotension with systolic blood pressure below 90 mmHg also demonstrated strong prognostic significance.

Reduced left ventricular ejection fraction below 35% independently predicted poor outcome, likely reflecting severe myocardial dysfunction and impaired compensatory reserve. Chronic kidney disease and multivessel coronary artery disease also remained statistically significant predictors after multivariable adjustment.

Interestingly, age alone demonstrated weaker independent prognostic impact after adjustment for hemodynamic variables, suggesting that physiologic instability may provide stronger short-term prognostic information than chronological age in emergency cardiovascular populations.

**Table 3.**
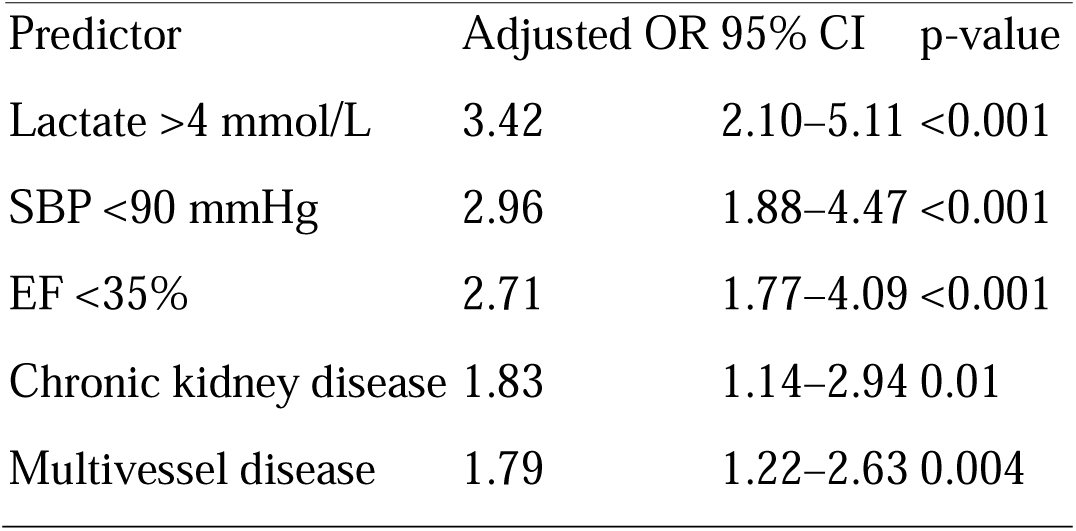
Independent Predictors of Major Adverse Cardiovascular Events.

| Predictor | Adjusted OR | 95% CI | p-value |
| --- | --- | --- | --- |
| Lactate >4 mmol/L | 3.42 | 2.10–5.11 | <0.001 |
| SBP <90 mmHg | 2.96 | 1.88–4.47 | <0.001 |
| EF <35% | 2.71 | 1.77–4.09 | <0.001 |
| Chronic kidney disease | 1.83 | 1.14–2.94 | 0.01 |
| Multivessel disease | 1.79 | 1.22–2.63 | 0.004 |

### Kaplan–Meier Survival Analysis

Kaplan–Meier survival analysis demonstrated significant early divergence between stable and unstable patient groups within the first 72 hours following emergency department admission. Mortality accumulation occurred most rapidly during the initial hospitalization period, particularly among patients requiring vasopressor therapy and mechanical ventilation.

At 30-day follow-up, survival probability remained significantly lower among unstable patients compared with stable patients. The observed survival separation suggests that early emergency hemodynamic deterioration has persistent prognostic implications extending beyond immediate coronary reperfusion.

The survival patterns observed in the present cohort are consistent with prior cardiovascular shock literature demonstrating that early physiologic compromise frequently initiates progressive inflammatory activation, ventricular remodeling, and multiorgan dysfunction even after successful coronary intervention (Reynolds & Hochman, 2008; Thiele et al., 2019).

**Figure 3.**
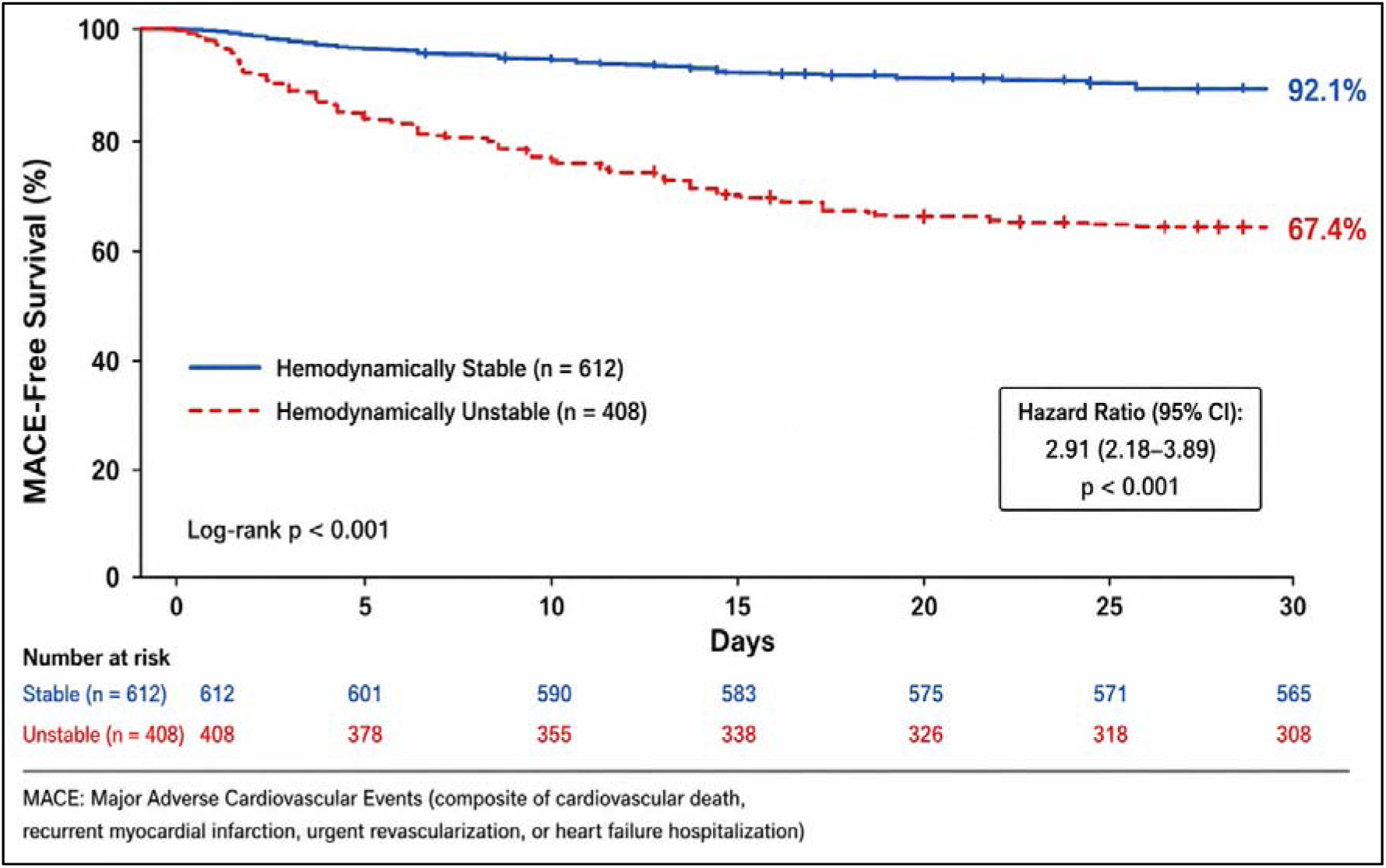
Kaplan–Meier Survival Curve

The Kaplan–Meier survival curve demonstrated early divergence between stable and unstable patient groups within the first 72 hours following emergency department presentation. Hemodynamically unstable patients experienced significantly lower short-term survival probability.

## Discussion

The present study demonstrated a strong association between early hemodynamic instability and adverse cardiovascular outcomes among patients presenting with acute coronary syndrome in a tertiary emergency cardiovascular center. Patients classified as hemodynamically unstable experienced significantly higher mortality, cardiogenic shock incidence, ventricular arrhythmia burden, ICU admission frequency, and composite major adverse cardiovascular events compared with stable patients.

These findings are clinically important because they reinforce the critical role of integrated emergency physiologic assessment during the earliest phase of ACS management. Traditional emergency cardiovascular evaluation frequently emphasizes electrocardiographic interpretation and biomarker confirmation; however, the present data suggest that dynamic hemodynamic profiling may provide equally important prognostic information.

One of the most notable findings in the current study involved the prognostic significance of elevated serum lactate concentration, which has similarly been associated with circulatory failure and increased cardiovascular mortality in previous emergency cardiovascular investigations (Mebazaa et al., 2015; Jentzer et al., 2019). Patients demonstrating lactate levels above 4 mmol/L experienced markedly increased adverse cardiovascular event rates even after adjustment for comorbid conditions and coronary anatomy. Elevated lactate likely reflects systemic tissue hypoperfusion, impaired oxygen delivery, neurohormonal activation, and evolving circulatory failure. Previous investigations have similarly identified lactate elevation as a predictor of mortality among critically ill cardiovascular patients.

Reduced left ventricular ejection fraction also emerged as an independent predictor of adverse outcomes. Severe ventricular dysfunction contributes to reduced cardiac output, impaired coronary perfusion reserve, pulmonary congestion, and progressive end-organ hypoperfusion. Importantly, bedside echocardiography now permits rapid emergency evaluation of ventricular performance and may therefore represent a valuable triage tool in emergency cardiovascular practice.

Another clinically relevant observation involved the significantly increased prevalence of multivessel coronary artery disease among unstable patients. Extensive coronary disease likely reflects larger ischemic burden, impaired collateral circulation, and greater susceptibility to cardiogenic shock. Contemporary cardiovascular intervention strategies increasingly support complete or staged revascularization among selected high-risk patients with multivessel disease.

The present findings are consistent with prior cardiovascular registries demonstrating that early physiologic deterioration strongly predicts adverse outcomes after myocardial infarction. Nevertheless, the current analysis contributes additional evidence by integrating multiple emergency hemodynamic variables rather than relying exclusively on isolated blood pressure measurements or Killip classification alone.

The study additionally highlights the evolving role of emergency cardiovascular critical care systems. Increasing integration of point-of-care ultrasonography, invasive arterial monitoring, bedside echocardiography, mechanical circulatory support, and advanced coronary intervention may improve survival among high-risk ACS populations. Future research should evaluate whether standardized hemodynamic assessment protocols can further optimize emergency cardiovascular outcomes.

Several limitations should be acknowledged. First, the retrospective design introduces potential selection bias and residual confounding despite multivariable adjustment. Second, the study was conducted within a tertiary referral center, potentially limiting external generalizability to smaller institutions. Third, serial hemodynamic changes after ICU admission were not comprehensively analyzed. Fourth, biomarker variability and treatment heterogeneity may have influenced outcomes.

Despite these limitations, the study provides clinically relevant evidence supporting the prognostic significance of early hemodynamic instability among ACS patients. Future prospective multicenter studies incorporating advanced hemodynamic monitoring and machine-learning-assisted risk stratification may further improve cardiovascular emergency triage systems.

## Conclusion

The present study demonstrated that early hemodynamic instability among patients presenting with acute coronary syndrome represents one of the strongest determinants of short-term cardiovascular morbidity and mortality in contemporary emergency cardiovascular practice. Patients who developed physiologic deterioration during the earliest phase of emergency department evaluation experienced substantially higher rates of cardiogenic shock, malignant ventricular arrhythmia, respiratory failure, acute kidney injury, intensive care utilization, and 30-day major adverse cardiovascular events compared with hemodynamically stable patients. Importantly, the prognostic impact of hemodynamic instability remained significant even after adjustment for baseline cardiovascular risk factors, coronary anatomy, and procedural characteristics, emphasizing that early circulatory compromise itself reflects a critical stage of cardiovascular decompensation rather than merely a secondary marker of disease severity. Among the evaluated hemodynamic parameters, elevated serum lactate concentration, severe hypotension, and markedly reduced left ventricular ejection fraction emerged as the most powerful independent predictors of adverse cardiovascular outcomes. These findings reinforce the concept that systemic hypoperfusion and impaired myocardial reserve are central pathophysiological mechanisms driving progression toward multiorgan dysfunction and cardiovascular collapse in ACS populations. The observation that survival curve divergence occurred within the first 72 hours following admission further highlights the importance of immediate physiologic assessment during the earliest stages of emergency cardiovascular management. The current analysis additionally demonstrated that hemodynamically unstable patients frequently presented with more extensive ischemic burden, including proximal left anterior descending artery occlusion, multivessel coronary artery disease, severe thrombotic lesions, and larger myocardial injury reflected by markedly elevated troponin concentrations. These angiographic and biochemical findings suggest that early hemodynamic deterioration is closely associated with extensive coronary compromise and advanced myocardial dysfunction. Consequently, integrated emergency hemodynamic assessment may serve not only as a prognostic marker but also as an indirect indicator of underlying coronary complexity and ischemic severity. From a clinical perspective, the findings of this study strongly support incorporation of comprehensive hemodynamic profiling into routine emergency cardiovascular triage systems. Contemporary ACS management has traditionally focused on electrocardiographic interpretation and rapid coronary reperfusion; however, the present results suggest that dynamic physiologic assessment provides equally important prognostic information capable of identifying high-risk patients before overt cardiogenic shock becomes clinically apparent. Early recognition of subtle hemodynamic deterioration may therefore permit more aggressive monitoring strategies, earlier intensive care escalation, optimized vasopressor utilization, timely mechanical circulatory support consideration, and improved allocation of cardiovascular critical care resources. The study also carries important implications for future cardiovascular emergency medicine research. As emergency departments increasingly adopt bedside echocardiography, point-of-care ultrasonography, invasive arterial monitoring, automated hemodynamic systems, and artificial intelligence–assisted predictive analytics, integration of multidimensional physiologic monitoring may substantially improve risk stratification accuracy among ACS populations. Future multicenter prospective investigations should evaluate whether standardized hemodynamic scoring systems combined with advanced cardiovascular imaging and biomarker analysis can further reduce mortality and improve long-term cardiovascular outcomes. Despite several limitations inherent to its retrospective single-center design, the present study provides clinically meaningful evidence emphasizing that early hemodynamic instability is not merely a transient physiologic abnormality but rather a powerful manifestation of advanced cardiovascular compromise associated with significantly increased mortality risk. The findings underscore the necessity of rapid identification, comprehensive physiologic monitoring, and early aggressive cardiovascular intervention among unstable ACS patients. Ultimately, the present study highlights that the first hours following emergency department presentation represent a critical therapeutic window during which timely recognition of hemodynamic deterioration may profoundly influence cardiovascular survival. Integration of advanced hemodynamic assessment into routine ACS management pathways may therefore represent an essential step toward improving emergency cardiovascular outcomes in high-risk myocardial infarction populations.

## Data Availability

All data produced in the present study are available upon reasonable request to the authors

## References

1. Amsterdam EA, Wenger NK, Brindis RG, Casey DE Jr, Ganiats TG, Holmes DR Jr, et al. 2014 AHA/ACC guideline for the management of patients with non–ST-elevation acute coronary syndromes. Circulation. 2014;130(25):e344-e426.

2. Anderson JL, Morrow DA. Acute myocardial infarction. N Engl J Med. 2017;376(21):2053–2064.

3. Antman EM, Cohen M, Bernink PJLM, McCabe CH, Horacek T, Papuchis G, et al. The TIMI risk score for unstable angina/non–ST elevation MI. JAMA. 2000;284(7):835–842.

4. Antman EM, Anbe DT, Armstrong PW, Bates ER, Green LA, Hand M, et al. ACC/AHA guidelines for the management of patients with ST-elevation myocardial infarction. Circulation. 2004;110(9):e82–e292.

5. Babaev A, Frederick PD, Pasta DJ, Every N, Sichrovsky T, Hochman JS. Trends in management and outcomes of patients with acute myocardial infarction complicated by cardiogenic shock. JAMA. 2005;294(4):448–454.

6. Bagai A, Chen AY, Wang TY, Alexander KP, Thomas L, Ohman EM, et al. Long-term outcomes among older patients with non–ST-segment elevation myocardial infarction complicated by cardiogenic shock. Am Heart J. 2013;166(2):298–305.

7. Basir MB, Schreiber TL, Grines CL, Dixon SR, Moses JW, Maini BS, et al. Effect of early initiation of mechanical circulatory support on survival in cardiogenic shock. Am J Cardiol. 2017;119(6):845–851.

8. Basir MB, Kapur NK, Patel K, Salam MA, Schreiber T, Kaki A, et al. Improved outcomes associated with the use of shock protocols. Catheter Cardiovasc Interv. 2019;93(7):1173–1183.

9. Bhatt DL, Lopes RD, Harrington RA. Diagnosis and treatment of acute coronary syndromes. JAMA. 2022;327(7):662–675.

10. Bøtker HE, Kharbanda R, Schmidt MR, Bøttcher M, Kaltoft AK, Terkelsen CJ, et al. Remote ischaemic conditioning before hospital admission in patients with acute myocardial infarction. Lancet. 2010;375(9716):727–734.

11. Byrne RA, Rossello X, Coughlan JJ, Barbato E, Berry C, Chieffo A, et al. 2023 ESC Guidelines for the management of acute coronary syndromes. Eur Heart J. 2023;44(38):3720–3826.

12. Cannon CP, Weintraub WS, Demopoulos LA, Vicari R, Frey MJ, Lakkis N, et al. Comparison of early invasive and conservative strategies in patients with unstable coronary syndromes. N Engl J Med. 2001;344(25):1879–1887.

13. Collet JP, Thiele H, Barbato E, Barthélémy O, Bauersachs J, Bhatt DL, et al. 2020 ESC Guidelines for the management of acute coronary syndromes in patients presenting without persistent ST-segment elevation. Eur Heart J. 2021;42(14):1289-1367.

14. Damman P, van’t Hof AWJ, ten Berg JM, Jukema JW, Appelman Y, Liem AH, et al. 2015 ESC Guidelines for acute coronary syndromes without persistent ST-segment elevation: recommendations and clinical implications. Neth Heart J. 2017;25(3):181–185.

15. De Luca G, Suryapranata H, Ottervanger JP, Antman EM. Time delay to treatment and mortality in primary angioplasty for acute myocardial infarction. Circulation. 2004;109(10):1223–1225.

16. De Waha S, Fuernau G, Eitel I, Desch S, Thiele H. Long-term prognosis after extracorporeal life support in refractory cardiogenic shock. JACC Cardiovasc Interv. 2016;9(19):2035–2044.

17. Eitel I, Desch S, Fuernau G, Hildebrand L, Gutberlet M, Schuler G, et al. Prognostic significance and determinants of myocardial salvage assessed by cardiovascular magnetic resonance in acute reperfused myocardial infarction. J Am Coll Cardiol. 2010;55(22):2470–2479.

18. Fox KAA, Dabbous OH, Goldberg RJ, Pieper KS, Eagle KA, Van de Werf F, et al. Prediction of risk of death and myocardial infarction in the six months after presentation with acute coronary syndrome. BMJ. 2006;333(7578):1091.

19. Fox KAA, Goodman SG, Klein W, Brieger D, Steg PG, Dabbous O, et al. Management of acute coronary syndromes: variations in practice and outcome. Eur Heart J. 2002;23(15):1177–1189.

20. Garcia-Garcia HM, McFadden EP, Farb A, Mehran R, Stone GW, Spertus J, et al. Standardized end point definitions for coronary intervention trials. Eur Heart J. 2018;39(23):2192–2207.

21. Gershlick AH, Khan JN, Kelly DJ, Greenwood JP, Sasikaran T, Curzen N, et al. Randomized trial of complete versus lesion-only revascularization in STEMI and multivessel disease. J Am Coll Cardiol. 2015;65(10):963–972.

22. Giugliano RP, White JA, Bode C, Armstrong PW, Montalescot G, Lewis BS, et al. Early versus delayed, provisional eptifibatide in acute coronary syndromes. N Engl J Med. 2009;360(21):2176–2190.

23. Goldberg RJ, Spencer FA, Gore JM, Lessard D, Yarzebski J. Thirty-year trends in the incidence and hospital death rates associated with cardiogenic shock in patients with acute myocardial infarction. Circulation. 2009;119(9):1211–1219.

24. Granger CB, Goldberg RJ, Dabbous O, Pieper KS, Eagle KA, Cannon CP, et al. Predictors of hospital mortality in the global registry of acute coronary events. Arch Intern Med. 2003;163(19):2345–2353.

25. Hannan EL, Zhong Y, Jacobs AK, Holmes DR, Walford G, Venditti FJ, et al. Patients with cardiogenic shock undergoing percutaneous coronary intervention. JACC Cardiovasc Interv. 2016;9(4):341–351.

26. Hochman JS, Sleeper LA, Webb JG, Sanborn TA, White HD, Talley JD, et al. Early revascularization in acute myocardial infarction complicated by cardiogenic shock. N Engl J Med. 1999;341(9):625–634.

27. Hochman JS, Sleeper LA, White HD, Dzavik V, Wong SC, Menon V, et al. One-year survival following early revascularization for cardiogenic shock. JAMA. 2001;285(2):190–192.

28. Ibanez B, James S, Agewall S, Antunes MJ, Bucciarelli-Ducci C, Bueno H, et al. 2017 ESC Guidelines for the management of acute myocardial infarction in patients presenting with ST-segment elevation. Eur Heart J. 2018;39(2):119–177.

29. Jentzer JC, van Diepen S, Barsness GW, Henry TD, Menon V, Rihal CS, et al. Cardiogenic shock classification to predict mortality in the cardiac intensive care unit. J Am Coll Cardiol. 2019;74(17):2117–2128.

30. Kastrati A, Mehilli J, Neumann FJ, Dotzer F, ten Berg J, Bollwein H, et al. Abciximab in patients with acute coronary syndromes undergoing percutaneous coronary intervention. N Engl J Med. 2004;350(3):232–238.

31. Keeley EC, Boura JA, Grines CL. Primary angioplasty versus intravenous thrombolytic therapy for acute myocardial infarction. Lancet. 2003;361(9351):13–20.

32. Killip T, Kimball JT. Treatment of myocardial infarction in a coronary care unit. Am J Cardiol. 1967;20(4):457–464.

33. Klein LW, Shaw RE, Krone RJ, Brindis RG, Anderson HV, Block PC, et al. Mortality after emergent percutaneous coronary intervention in cardiogenic shock. J Am Coll Cardiol. 2005;45(8):1211–1219.

34. Kosuge M, Kimura K, Ishikawa T, Ebina T, Hibi K, Tsukahara K, et al. Differences between men and women in terms of clinical features of ST-segment elevation acute myocardial infarction. Circ J. 2006;70(3):222–226.

35. Lemkes JS, Janssens GN, van der Hoeven NW, Jewbali LSD, Dubois EA, Meuwissen M, et al. Coronary angiography after cardiac arrest without ST-segment elevation. N Engl J Med. 2019;380(15):1397–1407.

36. Lopes RD, Heizer G, Aronson R, Vora AN, Massaro T, Mehran R, et al. Antithrombotic therapy after acute coronary syndrome or PCI in atrial fibrillation. N Engl J Med. 2019;380(16):1509–1524.

37. Mehta SR, Wood DA, Storey RF, Mehran R, Bainey KR, Nguyen H, et al. Complete revascularization with multivessel PCI for myocardial infarction. N Engl J Med. 2019;381(15):1411–1421.

38. Mebazaa A, Yilmaz MB, Levy P, Ponikowski P, Peacock WF, Laribi S, et al. Recommendations on pre-hospital and early hospital management of acute heart failure. Eur J Heart Fail. 2015;17(6):544–558.

39. Menon V, Fincke R. Cardiogenic shock: a summary of the randomized SHOCK trial. Curr Opin Cardiol. 2003;18(5):360–365.

40. Mensah GA, Fuster V, Murray CJL, Roth GA. Global burden of cardiovascular diseases and risks, 1990-2022. J Am Coll Cardiol. 2023;82(25):2350–2473.

41. Montalescot G, Bolognese L, Dudek D, Goldstein P, Hamm C, Tanguay JF, et al. Pretreatment with prasugrel in non–ST-segment elevation acute coronary syndromes. N Engl J Med. 2013;369(11):999–1010.

42. Morrow DA, Antman EM, Charlesworth A, Cairns R, Murphy SA, de Lemos JA, et al. TIMI risk score for ST-elevation myocardial infarction. Circulation. 2000;102(17):2031–2037.

43. Morrow DA, Braunwald E. Future of biomarkers in acute coronary syndromes. Circulation. 2003;108(3):250–252.

44. Ndrepepa G, Kastrati A, Mehilli J, Neumann FJ, Ten Berg J, Bruskina O, et al. Mechanical reperfusion and long-term mortality in patients with acute myocardial infarction presenting with cardiogenic shock. JAMA. 2005;293(4):437–444.

45. Neumann FJ, Sousa-Uva M, Ahlsson A, Alfonso F, Banning AP, Benedetto U, et al. 2018 ESC/EACTS Guidelines on myocardial revascularization. Eur Heart J. 2019;40(2):87–165.

46. Neumann JT, Sörensen NA, Rübsamen N, Ojeda F, Renné T, Qaderi V, et al. Discrimination of patients with type 2 myocardial infarction. Eur Heart J. 2017;38(5):351–359.

47. O’Donoghue ML, Braunwald E, White HD, Steen DP, Lukas MA, Tarka E, et al. Effect of darapladib on major coronary events after an acute coronary syndrome. JAMA. 2014;312(10):1006–1015.

48. O’Gara PT, Kushner FG, Ascheim DD, Casey DE Jr, Chung MK, de Lemos JA, et al. 2013 ACCF/AHA guideline for the management of ST-elevation myocardial infarction. Circulation. 2013;127(4):e362-e425.

49. Park DW, Clare RM, Schulte PJ, Pieper KS, Shaw LK, Califf RM, et al. Extent, location, and clinical significance of non-infarct-related coronary artery disease among patients with STEMI. JAMA. 2014;312(19):2019–2027.

50. Patel MR, Calhoon JH, Dehmer GJ, Grantham JA, Maddox TM, Maron DJ, et al. ACC/AATS/AHA/ASE/ASNC/SCAI/SCCT/STS appropriate use criteria for coronary revascularization. J Am Coll Cardiol. 2017;69(17):2212–2241.

51. Puymirat E, Simon T, Cayla G, Cottin Y, Elbaz M, Coste P, et al. Acute myocardial infarction: changes in patient characteristics, management, and 6-month outcomes over a period of 20 years. Circulation. 2012;126(10):1110–1118.

52. Rathod KS, Koganti S, Iqbal MB, Jain AK, Kalra SS, Astroulakis Z, et al. Contemporary trends in cardiogenic shock. Int J Cardiol. 2018;260:7–13.

53. Reynolds HR, Hochman JS. Cardiogenic shock: current concepts and improving outcomes. Circulation. 2008;117(5):686–697.

54. Roe MT, Messenger JC, Weintraub WS, Cannon CP, Fonarow GC, Dai D, et al. Treatments, trends, and outcomes of acute myocardial infarction and percutaneous coronary intervention. J Am Coll Cardiol. 2010;56(4):254–263.

55. Roffi M, Patrono C, Collet JP, Mueller C, Valgimigli M, Andreotti F, et al. 2015 ESC Guidelines for the management of acute coronary syndromes in patients presenting without persistent ST-segment elevation. Eur Heart J. 2016;37(3):267–315.

56. Roth GA, Mensah GA, Johnson CO, Addolorato G, Ammirati E, Baddour LM, et al. Global burden of cardiovascular diseases and risk factors, 1990-2019. J Am Coll Cardiol. 2020;76(25):2982–3021.

57. Sabatine MS, Cannon CP, Gibson CM, López-Sendón JL, Montalescot G, Theroux P, et al. Addition of clopidogrel to aspirin and fibrinolytic therapy for myocardial infarction. N Engl J Med. 2005;352(12):1179–1189.

58. Sabatine MS, Giugliano RP, Keech AC, Honarpour N, Wiviott SD, Murphy SA, et al. Evolocumab and clinical outcomes in patients with cardiovascular disease. N Engl J Med. 2017;376(18):1713–1722.

59. Sanborn TA, Sleeper LA, Webb JG, French JK, Bergman G, Parikh M, et al. Correlates of one-year survival in patients with cardiogenic shock complicating acute myocardial infarction. J Am Coll Cardiol. 2003;42(8):1373–1379.

60. Schiele F, Gale CP, Bonnefoy E, Capuano F, Claeys MJ, Danchin N, et al. Quality indicators for acute myocardial infarction. Eur Heart J Acute Cardiovasc Care. 2017;6(1):34–59.

61. Shah RU, de Lemos JA, Wang TY, Chen AY, Thomas L, Sutton NR, et al. Post-hospital outcomes of patients with acute myocardial infarction with cardiogenic shock. J Am Coll Cardiol. 2016;67(7):739–747.

62. Shavadia JS, Zheng Y, Udell JA, Granger CB, Peterson ED, Pieper KS, et al. Predictors of long-term mortality in patients with cardiogenic shock complicating acute myocardial infarction. Am J Cardiol. 2016;117(3):343–349.

63. Sionis A, Rivas-Lasarte M, Mebazaa A, Tarvasmäki T, Sans-Roselló J, Tolppanen H, et al. Current use and impact on 30-day mortality of pulmonary artery catheter in cardiogenic shock patients. J Intensive Care Med. 2020;35(12):1426–1433.

64. Steg PG, James SK, Atar D, Badano LP, Blömstrom-Lundqvist C, Borger MA, et al. ESC Guidelines for the management of acute myocardial infarction in patients presenting with ST-segment elevation. Eur Heart J. 2012;33(20):2569–2619.

65. Stone GW, Lansky AJ, Pocock SJ, Gersh BJ, Dangas G, Wong SC, et al. Paclitaxel-eluting stents versus bare-metal stents in acute myocardial infarction. N Engl J Med. 2009;360(19):1946–1959.

66. Stone GW, Maehara A, Lansky AJ, de Bruyne B, Cristea E, Mintz GS, et al. A prospective natural-history study of coronary atherosclerosis. N Engl J Med. 2011;364(3):226–235.

67. Szummer K, Wallentin L, Lindhagen L, Alfredsson J, Erlinge D, Held C, et al. Improved outcomes in patients with ST-elevation myocardial infarction during the last 20 years. Eur Heart J. 2017;38(41):3056–3065.

68. Tarvasmäki T, Lassus J, Varpula M, Sionis A, Sund R, Køber L, et al. Current real-life use of vasopressors and inotropes in cardiogenic shock. Crit Care. 2016;20(1):208.

69. Thiele H, Akin I, Sandri M, Fuernau G, de Waha S, Meyer-Saraei R, et al. PCI strategies in patients with acute myocardial infarction and cardiogenic shock. N Engl J Med. 2017;377(25):2419–2432.

70. Thiele H, Ohman EM, de Waha-Thiele S, Zeymer U, Desch S. Management of cardiogenic shock complicating myocardial infarction. Eur Heart J. 2019;40(32):2671–2683.

71. Thiele H, Zeymer U, Neumann FJ, Ferenc M, Olbrich HG, Hausleiter J, et al. Intraaortic balloon support for myocardial infarction with cardiogenic shock. N Engl J Med. 2012;367(14):1287–1296.

72. Thygesen K, Alpert JS, Jaffe AS, Chaitman BR, Bax JJ, Morrow DA, et al. Fourth universal definition of myocardial infarction. Circulation. 2018;138(20):e618–e651.

73. Valgimigli M, Bueno H, Byrne RA, Collet JP, Costa F, Jeppsson A, et al. 2017 ESC focused update on dual antiplatelet therapy in coronary artery disease. Eur Heart J. 2018;39(3):213–260.

74. Van de Werf F, Bax J, Betriu A, Blomstrom-Lundqvist C, Crea F, Falk V, et al. Management of acute myocardial infarction in patients presenting with persistent ST-segment elevation. Eur Heart J. 2008;29(23):2909–2945.

75. van Diepen S, Katz JN, Albert NM, Henry TD, Jacobs AK, Kapur NK, et al. Contemporary management of cardiogenic shock. Circulation. 2017;136(16):e232–e268.

76. van Diepen S, Vavalle JP, Newby LK, Clare R, Pieper KS, Ezekowitz JA, et al. The systemic inflammatory response syndrome in patients with ST-segment elevation myocardial infarction. Crit Care Med. 2013;41(9):2080–2087.

77. Vahdatpour C, Collins D, Goldberg S. Cardiogenic shock. J Am Heart Assoc. 2019;8(8):e011991.

78. Vranckx P, Valgimigli M, Windecker S, Steg PG, Hamm C, Juni P, et al. Long-term ticagrelor monotherapy versus standard dual antiplatelet therapy after PCI. Lancet. 2018;392(10151):940–949.

79. Wallentin L, Becker RC, Budaj A, Cannon CP, Emanuelsson H, Held C, et al. Ticagrelor versus clopidogrel in patients with acute coronary syndromes. N Engl J Med. 2009;361(11):1045–1057.

80. Widimský P, Wijns W, Fajadet J, de Belder M, Knot J, Aaberge L, et al. Reperfusion therapy for ST-elevation acute myocardial infarction in Europe. Eur Heart J. 2010;31(8):943–957.

81. Windecker S, Kolh P, Alfonso F, Collet JP, Cremer J, Falk V, et al. 2014 ESC/EACTS Guidelines on myocardial revascularization. Eur Heart J. 2014;35(37):2541–2619.

82. Wiviott SD, Braunwald E, McCabe CH, Montalescot G, Ruzyllo W, Gottlieb S, et al. Prasugrel versus clopidogrel in patients with acute coronary syndromes. N Engl J Med. 2007;357(20):2001–2015.

83. Yeh RW, Sidney S, Chandra M, Sorel M, Selby JV, Go AS. Population trends in the incidence and outcomes of acute myocardial infarction. N Engl J Med. 2010;362(23):2155–2165.

84. Zeymer U, Hochadel M, Hauptmann KE, Wiegand K, Schuhmacher B, Brachmann J, et al. Intra-aortic balloon pump in patients with acute myocardial infarction complicated by cardiogenic shock. Clin Res Cardiol. 2013;102(3):223–227.

85. Zeymer U, Bueno H, Granger CB, Hochman J, Huber K, Lettino M, et al. Acute cardiovascular care association position statement for the diagnosis and treatment of patients with acute myocardial infarction complicated by cardiogenic shock. Eur Heart J Acute Cardiovasc Care. 2020;9(2):183–197.

86. Zeymer U, Freund A, Hochadel M, Ostad MA, Belohlavek J, Rokyta R, et al. Venoarterial extracorporeal membrane oxygenation in patients with infarct-related cardiogenic shock. Circulation. 2023;147(6):454–464.

87. Zijlstra F, Hoorntje JCA, de Boer MJ, Reiffers S, Miedema K, Ottervanger JP, et al. Long-term benefit of primary angioplasty as compared with thrombolytic therapy for acute myocardial infarction. N Engl J Med. 1999;341(19):1413–1419.

88. Zimarino M, Curzen N, Cicchitti V, De Caterina R. The adequacy of myocardial revascularization in patients with multivessel coronary artery disease. Int J Cardiol. 2013;168(3):1748–1757.

89. Zipes DP, Camm AJ, Borggrefe M, Buxton AE, Chaitman B, Fromer M, et al. ACC/AHA/ESC guidelines for management of patients with ventricular arrhythmias and prevention of sudden cardiac death. Circulation. 2006;114(10):e385–e484.

